# Substance Use and Overdose in Pennsylvania Libraries, 2019

**DOI:** 10.1101/2024.01.03.24300786

**Authors:** Rachel Feuerstein-Simon, Margaret Lowenstein, Claire Szapary, Omaya Torres, Abby Dolan, Aminata Jalloh, Zachary F. Meisel, Carolyn C. Cannuscio

## Abstract

This study examines the implementation of naloxone distribution initiatives in Pennsylvania public libraries following a nationwide program offering free Narcan. We conducted a cross-sectional telephone survey with a random sample of Pennsylvania public libraries (n=352). Overall, nearly one-quarter of respondents reported stocking naloxone (23.9%), and over one in ten libraries reported a previous on-site overdose (11.9%). Nearly 30% of respondents had received naloxone training. Significant predictors of on-site overdoses included the library’s county urbanization status and higher county-level overdose mortality rates. Among libraries that stocked naloxone, 73% obtained the medication from local health departments or community-based organizations. This study underscores the role of public libraries in opioid overdose crisis response and the need for tailored strategies to enhance naloxone accessibility, especially in high-risk urban areas. Collaboration between libraries, public health entities, and pharmaceutical companies is crucial to amplify naloxone distribution efforts and address the escalating opioid overdose crisis.

## Introduction

Overdose deaths are surging in the United States, with more than 100,000 overdose fatalities recorded in 2021 alone.^1^ This represents a staggering 160% increase over the decade spanning from 2011 to 2021, largely driven by the proliferation of fentanyl and other synthetic opioids.^1,2^ A vital strategy in addressing the overdose crisis is wider availability of the opioid overdose antidote, naloxone.^3–5^ Research confirms the safe use of naloxone by both first responders and laypeople, and community-based overdose education and naloxone distribution (OEND) programs have demonstrated effectiveness in reducing overdose mortality.^4,5^ However, current naloxone distribution strategies are insufficient in the face of escalating overdose deaths.

Given the magnitude of the crisis, there is an urgent need to expand overdose prevention efforts beyond traditional groups like first responders and people with opioid use disorder (OUD). Public libraries have emerged as promising sites for harm reduction interventions and overdose response efforts.^6–9^ Public libraries host over one billion annual visits and serve as information hubs and safety nets for vulnerable populations.^10^ Library staff report routinely answering patron concerns regarding health, and prior research has shown that approximately half of library staff specifically offered mental health and substance use-related assistance in the prior month.^9^ Public libraries have also become locations of on-site drug use and overdose. Evidence suggests that approximately 12% of US public libraries have had an on-site overdose in the prior year.^8,9^

In October 2018, the manufacturer of the naloxone nasal spray, Narcan, launched a nationwide initiative offering free Narcan to any school or library willing to stock the medication.^11^ The aim of this study was to assess naloxone uptake in Pennsylvania public libraries, thereby tracking the success of the manufacturer distribution effort. The study also evaluated the one-year cumulative incidence of overdose in Pennsylvania public libraries, as well as predictors of both naloxone uptake and on-site overdose.

## Methods

### Study design and sample

This study employed a cross-sectional telephone survey design. Survey respondents were recruited from a 70% random sample of all library branches in Pennsylvania documented in the 2016 Public Library Survey Census. Of 643 library branches across Pennsylvania, 446 were randomly selected for study recruitment. One person was invited to respond from each library branch.

### Survey design and administration

The brief survey focused on two queries: 1) whether the library currently stocked naloxone, and 2) if there had been an on-site overdose. Additional questions asked about interactions with patrons regarding substance use and addiction and its treatment. Library staff who reported that their library stocked naloxone on-site were asked questions about how the medication had been obtained. Libraries that had experienced an on-site overdose were asked additional questions regarding response to the incident.

### Data collection and sources

Between June-August 2019, survey data were collected by five research assistants (RA) trained in qualitative methods, including interview techniques. RAs were instructed to request to speak with library directors/branch managers, or a designated person with knowledge of naloxone/on-site overdose. Libraries were considered non-responders if we were unable to reach them after three telephone attempts at different times of day. All data were documented and stored in REDCap, including verbal participation consent.

We used publicly available datasets for the analysis of county-level predictors of overdose and naloxone uptake. We used the Centers for Disease Control and Prevention (CDC) Wide-ranging Online Data for Epidemiologic Research (WONDER) database to ascertain 2018 county-level overdose mortality. Each library was classified as metropolitan (metro) or nonmetropolitan (nonmetro) based on county-level data from Economic Research Service (ERS) Rural-Urban Continuum Codes. To assess county-level socioeconomic factors, we use the Small Area Income and Poverty Estimates (SAIPE) State and County Estimates for 2018. To collect library-level characteristics, we used the Institute of Museum and Library Services (IMLS) Public Libraries Survey (PLS).

### Analysis

The survey data were analyzed using R version 4.3.1. Descriptive statistics were calculated to characterize institution and community-level features of the responding libraries. We used logistic regression to assess predictors of naloxone uptake and overdose. This study was approved by the University of Pennsylvania Institutional Review Board.

## Results

A total of n=352 respondents, each representing a different library branch, completed the telephone survey (response rate=79.0%). Table 1 presents library and community-level characteristics of the responding libraries. A majority of responding libraries were located in counties classified as metropolitan (77.6%) and had household incomes below the US median (55.1%). The median county overdose death rate was 36.1 deaths per 100,000 population per year. Over one in ten libraries reported a previous on-site overdose, and a minority of libraries stocked naloxone (23.9%).

**Table 1.** Institution and county-level characteristics of responding libraries.

|  | **Total**<br>**(N=352)** |
| --- | --- |
| **County population** |  |
| Mean (SD) | 466000 (465000) |
| Median [Min, Max] | 277000 [4490, 1580000] |
| **County urbanization classification** |  |
| Metro | 273 (77.6%) |
| Non-metro | 79 (22.4%) |
| **County median household income** |  |
| Above US median household income | 156 (44.3%) |
| Below US median household income | 194 (55.1%) |
| **Proportion of county living in poverty** |  |
| Mean (SD) | 11.9 (3.63) |
| Median [Min, Max] | 11.3 [6.50, 22.3] |
| **County overdose death rate** |  |
| Mean (SD) | 34.1 (12.5) |
| Median [Min, Max] | 33.9 [0, 61.1] |
| **Per capita library expenditures** |  |
| Mean (SD) | 33.9 (36.7) |
| Median [Min, Max] | 21.8 [4.49, 201] |
| **Previous on-site overdose** |  |
| No | 310 (88.1%) |
| Yes | 42 (11.9%) |
| **Naloxone available at library** |  |
| No | 267 (75.9%) |
| Yes | 84 (23.9%) |
| **Library staff trained to use naloxone** |  |
| No | 236 (67.0%) |
| Yes | 100 (28.4%) |

Table 2 shows correlates of having had an on-site overdose among participating libraries. The strongest predictor of on-site overdose was county urbanization, with libraries located in metropolitan areas over four times as likely as those in non-metro areas to have had an on-site overdose. Libraries in counties with median overdose death rates above the Pennsylvania median overdose death rate were more than three times as likely as those in lower-overdose-mortality counties to have had an on-site overdose, a difference that was statistically significant. There was no significant association between county-level household income and on-site overdose.

**Table 2.** Predictors of on-site overdose.

| Predictor | OR | 95% CI | p |
| --- | --- | --- | --- |
| Metropolitan county | 4.22 | 1.48-17.8 | <0.05 |
| County overdose death rate above PA median | 3.38 | 1.68-7.30 | <0.01 |
| Median household income below US median | 0.825 | 0.43-1.59 | >0.05 |

Table 3 shows predictors of naloxone uptake in participating public libraries. Similar to on-site overdose, the strongest correlate of naloxone uptake was county urbanization, with libraries located in metropolitan areas nearly eight times as likely to stock naloxone as those in non-metropolitan areas. Further, counties with higher overdose death rates were more likely to stock naloxone. Specifically, libraries in counties with overdose death rates above the Pennsylvania median were 1.94 times as likely to stock naloxone, compared to libraries in counties with overdose death rates below the state median.

**Table 3.** Predictors of naloxone uptake.

| Predictor | OR | 95% CI | p |
| --- | --- | --- | --- |
| Metropolitan county | 7.81 | 3.11-26.3 | <0.001 |
| County overdose death rate above PA median | 1.94 | 1.17-3.23 | <0.05 |
| Median household income below US median | 1.25 | 0.765-2.08 | >0.05 |
| Had a prior on-site overdose | 3.10 | 1.58-6.04 | <0.001 |

Forty six percent of libraries that reported having a prior on-site overdose reported stocking naloxone. Among those libraries who stocked naloxone on-site (23.9%), the vast majority (73%) reported acquiring the medication either from a local health department or community-based organization. Just over one percent of libraries acquired naloxone from the pharmaceutical company or purchased it themselves. Compared to the proportion stocking naloxone on-site, a slightly higher proportion of respondents had been trained to use naloxone (28.4%). Among the library staff trained to use naloxone, nearly 90% attended an in-person training.

## Discussion

As the US responds to the deadly opioid overdose epidemic, we need multiple strategies both to prevent overdoses and to respond rapidly when they occur. Findings from this study reinforce prior reports that between 10-17% of libraries had experienced an on-site overdose in a one-year period.^8,9^ Given that drug use and overdose occur in public libraries, institutional responses must become part of our public health approach to preventing overdose deaths.

Public health professionals need better tools for predicting when and where overdoses will happen, so that institutions and individuals can prepare and intervene promptly to save lives. Beyond the finding that metropolitan area libraries are at increased risk for on-site overdose, what else predicts higher-risk sites or times? Does library location (e.g., near transit hubs or drug markets), design (e.g., open layout vs. other), or policy (e.g., bathroom time limits; hours of operation) predict on-site drug use or overdose? By identifying predictors of drug use or overdose on-site, public health researchers can assist institutions like libraries in recognizing and preparing for or remedying risks. In addition, by learning more about why people use drugs in public places, we can better design supports for people who use drugs. Anderson and colleagues have reported that people who use drugs prefer not to use in places frequented by children, for example--thus people may use places like libraries as a last resort.^12^ Given that library staff have expressed concern about being seen or frequented as de facto supervised injection sites^7^, the data from this paper could be introduced in public conversations about designated overdose prevention facilities.

This paper raised important questions regarding why libraries did not take advantage of the Narcan manufacturer program to distribute the drug for free. The majority of libraries with naloxone had received it through community organizations or health departments. Less than 2% of libraries in this PA survey had received the drug for free from the manufacturer. Evidence from this study comports with findings from media reports that suggest 2.2% of all US public libraries accepted the free doses, and another multi-state survey that showed that only 1.2% of libraries with naloxone had acquired it through the manufacturer initiative.^8,13^ Was this low rate of uptake due to preferences of libraries (or their leaders), or was it a result of specific manufacturer program features, such as onerous paperwork? Given the potential lifesaving benefits of naloxone, the reasons for the manufacturer’s programmatic failure should be explored further. Such findings would not only illuminate how better to help libraries prepare for overdoses, but would also offer insights potentially relevant to other institutions, like schools or workplaces, that could stock naloxone for emergencies. Additionally, very few libraries purchased naloxone from pharmacies, despite the widespread availability of the drug under standing orders (prior to the recent shift to over-the-counter status). Was there a shortage of time, money, will, or some other resource necessary for libraries to overcome procurement barriers? How will this shift now that naloxone is available over-the-counter? How can libraries (and other public institutions) best be supported, so that organizations and their staff are prepared for rapid response when a person overdoses on site?

While this paper focuses on overdoses in public libraries, we can think of libraries as a model for other public places where drug use and overdose may occur. By identifying high-risk public sites, and by preparing institutions and organizations with naloxone and overdose reversal training, the burden of overdose prevention and response shifts from people who use drugs--who are disproportionately vulnerable--to the community. This move toward place-based preventive responses and community care must become a pillar of our country’s public health approach to saving lives.

## Data Availability

A limited dataset produced in the present study is available upon reasonable request to the authors

## Notes

### Competing Interest Statement

The authors have declared no competing interest.

### Funding Statement

This study did not receive any funding

### Author Declarations

The Institutional Review Board University of Pennsylvania

## CITATIONS

1. Centers for Disease Control and Prevention. Drug Overdose Death Rates. Published June 30, 2023. https://nida.nih.gov/research-topics/trends-statistics/overdose-death-rates

2. Centers for Disease Control and Prevention. Drug Overdose Mortality by State. Published March 1, 2022. https://www.cdc.gov/nchs/pressroom/sosmap/drug_poisoning_mortality/drug_poisoning.htm

3. Pitt AL, Humphreys K, Brandeau ML. Modeling Health Benefits and Harms of Public Policy Responses to the US Opioid Epidemic. Am J Public Health. 2018;108(10):1394–1400. doi:10.2105/AJPH.2018.304590

4. Walley AY, Xuan Z, Hackman HH, et al. Opioid overdose rates and implementation of overdose education and nasal naloxone distribution in Massachusetts: interrupted time series analysis. BMJ. 2013;346(jan30 5):f174–f174. doi:10.1136/bmj.f174

5. Davis CS, Ruiz S, Glynn P, Picariello G, Walley AY. Expanded Access to Naloxone Among Firefighters, Police Officers, and Emergency Medical Technicians in Massachusetts. Am J Public Health. 2014;104(8):e7–e9. doi:10.2105/AJPH.2014.302062

6. Lowenstein M, Feuerstein-Simon R, Dupuis R, et al. Overdose Awareness and Reversal Trainings at Philadelphia Public Libraries. Am J Health Promot. 2021;35(2):250–254. doi:10.1177/0890117120937909

7. Lowenstein M, Feuerstein-Simon R, Sheni R, et al. Public Libraries as Partners in Confronting the Overdose Crisis: A Qualitative Analysis. Subst Abuse. 2021;42(3):302–309. doi:10.1080/08897077.2019.1691129

8. Feuerstein-Simon R, Lowenstein M, Dupuis R, et al. Substance Use and Overdose in Public Libraries: Results from a Five-State Survey in the US. J Community Health. 2022;47(2):344–350. doi:10.1007/s10900-021-01048-2

9. Whiteman ED, Dupuis R, Morgan AU, et al. Public Libraries As Partners for Health. Prev Chronic Dis. 2018;15:170392. doi:10.5888/pcd15.170392

10. Shubik-Richards C, Dowdall E. The Library in the City: Changing Demands and a Challenging Future. The Pew Charitable Trusts Philadelphia Research Initiative; 2012.

11. Inklebarger T. Company to Supply Free Narcan to Libraries. Am Libr. Published online October 24, 2018. https://americanlibrariesmagazine.org/blogs/the-scoop/narcan-company-supply-free-narcan-to-libraries/

12. Harris RE, Richardson J, Frasso R, Anderson ED. Perceptions about supervised injection facilities among people who inject drugs in Philadelphia. Int J Drug Policy. 2018;52:56–61. doi:10.1016/j.drugpo.2017.11.005

13. Anne Ford. Narcan or No? Am Libr Mag. Published online June 1, 2020. Accessed September 23, 2023. https://americanlibrariesmagazine.org/2020/06/01/narcan-or-no-libraries-naloxone/

